# Patients Accessing their Online Records in England: A Survey of General Practitioners’ Experiences and Opinions

**DOI:** 10.1101/2023.08.21.23294326

**Authors:** Charlotte Blease, Anna Kharko, Zhiyong Dong, Ray Jones, Gail Davidge, Maria Hägglund, Andrew Turner, Catherine M. DesRoches, Brian McMillan

## Abstract

**Objective:** To describe the experiences and opinions of general practitioners (GPs) in England regarding patients having access to their full online GP health record.

**Design:** Convenience sample, online survey.

**Participants:** 400 registered GPs in England.

**Main outcome measures:** Investigators measured GPs’ experiences and opinions about online record access (ORA) including on patient care, and on their practice.

**Results:** A total of 400 GPs from all regions of England responded. A minority (130, 33%) believed ORA was a good idea. Most GPs believed a majority of patients would worry more (364, 91%) or find their GP records more confusing than helpful (338, 85%). In contrast, most GPs believed a majority of patients would find significant errors in their records (240, 60%), would better remember their care plan (280, 70%), and feel more in control of their care (243, 60%). The majority believed they will/already spend more time addressing patients’ questions outside of consultations (357, 89%), that consultations will/already take significantly longer (322, 81%), and reported they will be/already are less candid in their documentation (289, 72%) after ORA. Nearly two thirds of GPs believed ORA would increase their litigation (246, 62%).

**Conclusions:** Similar to clinicians in other countries, GPs in our sample were sceptical of ORA believing patients would worry more and find their records more confusing than helpful. Most GPs also believed the practice would exacerbate work burdens. However, the majority of GPs in this survey also agreed there were multiple benefits to patients having online access to their primary care health record.

## Introduction

In 2021, the National Health Service England (NHSE) announced plans that patients 16 and over would have prospective access to their primary care record online, by default.[1] Although these plans have not yet been fully implemented, by March 2023, one in 5 English primary care practices switched on this functionality, enabling - at least in theory - 6.5 million patients to see new information added to their record using online services such as the National Health Service (NHS) App[2] Access includes test and lab results, secondary care letters, lists of medications, and the free text written by general practitioners (GPs) during consultations. Since April 2019, the GP contract in England already committed practices to offer new patients full prospective online access to their record: however, this was widely interpreted to mean access would be granted only after a patient request to GPs.[3] The new NHS England announcement specified that access would be enabled automatically and by default: that is, without requiring patients to submit a request for access. On 6 March 2023, NHSE announced a new GP contract which will impose ORA by default by October 2023[4].

The UK is comprised of publicly funded healthcare systems across each of its four countries: NHSE, NHS Scotland, NHS Wales and Health and Social Care in Northern Ireland. Within the UK, NHSE is the furthest along with respect to implementing ORA. England is not the first nor only country to implement default patient access to electronic health records. In some countries, such as the Nordic countries and the U.S., the practice is advanced.[5] For example, between 2012 to 2018, all patients in Sweden obtained access to their electronic records.[6] By 2021 in the U.S., 55 million people were already offered online access to their free text entries written by clinicians – a practice commonly referred to as “open notes.” Starting April 2021, new U.S. federal rules mandated, with few permitted exemptions, all patients be offered rapid access to their full electronic record, including open notes, without charge.[7,8] Although patients often welcome transparency, studies show many doctors, especially those without experience of the practice express scepticism about patient access including open notes.[9–13]

A literature review of primary care electronic health records in the United Kingdom is available elsewhere [14]. Less is known about the experiences and opinions of doctors in the UK regarding patients having access to their health record online.[14] Recently, however, researchers have begun to explore doctors’ views.[15] In 2022, Turner et al. conducted a qualitative study including 16 general practice staff in England who had experience of the practice. Participants expressed concerns that access negatively affected quality of record entries, patient safety, and workload.[16] Another study by Louch et al. explored the views of 19 primary care staff involved in a variety of clinical and non-clinical roles and found respondents to be generally supportive of patient online record access but were uncertain about the impact on patient-clinician relationships and patient safeguarding.[17] To date, these findings echo multiple studies published in Sweden and the U.S. where clinicians also express worries related to potential harms but also benefits of patient online record access.[9–13] Moreover, research in England exploring patients’ experiences echo findings in other countries,[18] showing that patients who access their records derive many benefits including feeling more in control of their care.[19,20] Although a growing number of small qualitative studies have explored the views of primary care staff in England about patient online record access (hereafter ‘ORA’), research in this area is limited. In this study, our aim was to address this gap by sampling a larger number of registered GPs in England to explore their general experiences and opinions about the potential impact of ORA on both patients and GPs.

## Methods

### Study population

Participants in this survey were sampled from GPs in England registered with the clinician marketing service Doctors.net.uk. This is the largest professional network and online information service of UK doctors with 248,326 doctors out of a total of 355,250 UK doctors (70%) registered with it. Approximately, 21,250 GPs out of a total of 36,752 registered and working in the UK (58%) are active in the community in any 90-day period. Ethical approval for the study was obtained from Beth Israel Deaconess Medical Center, Harvard Medical School (Protocol # 2021P000626). At Doctors.net.uk, a percentage of GPs active within the community consent to being sent survey invitations via email: this percentage differs according to those who are active in any given period. Therefore, depending on how GPs consented to receive survey invitations, our study was advertised via email or displayed on the Doctors.net.uk home pages of a sample of GPs between 10th and 31st March 2022. We asked Doctors.net.uk to invite a random sample stratified by gender and age using demographic information about currently registered GPs working in England provided by the General Medical Council (GMC) the GMC Data Explorer (https://data.gmc-uk.org/gmcdata/home/#/). We have obtained samples from Doctors.net.uk in previous studies using similar methods.[21] Doctors.net.uk invited 720 GPs by email and also by invitations embedded in their Doctors.net.uk homepages; a further 2,072 GPs were invited to participate only via links on their homepages.

All invited GPs were assured that their identities would not be disclosed to investigators, and participants gave informed consent before taking part in the survey. A small incentive of ‘1000 eSR’ points worth £7.50 ($8.80, €8.83) in exchangeable shopping vouchers was provided on completion, and participants were required to respond to every closed ended question to complete the survey. Further questions were embedded within the survey to determine whether respondents were currently practising as GPs in England. Data collection terminated when we received 400 completed survey responses.

### Survey instrument

The study team adapted a Survey instrument originally developed to explore US primary care physicians’ views and experiences with open notes.[10] We modified and shortened the survey in consultation with GPs in England and piloted the survey with GP colleagues in the UK (*n* = 5) to ensure face validity. The survey was timed to take around 5 minutes to complete.

The survey was divided into two main sections (see Supplement 1). Part One examined the impact of ORA on patients, and opened with the statement, “The following questions ask for your understanding, experiences, and opinions about offering patients full online access to their GP health records including the potential impact on patients’ care. By ‘full online access’ we mean all information on the electronic record from the date the patient requested access that is visible to GPs including the patient’s allergies, immunizations, letters, medication lists, test/lab results, problem lists, and the free text comments written by clinicians.” Next, using seven scalar options (including ‘none’) participants were requested to estimate what percentage of their patients were currently offered full online access to their records. Participants were also asked, “If your patients were/are offered access to your free text comments online, how many patients do you estimate would read them?” GPs were invited to respond using six scalar percentage options. A third set of questions stated, “We are interested in your opinions about the effect on patients of reading GP health records online, even if none of your patients have requested access. Please indicate how strongly you agree or disagree with the statements below.” GPs were invited to offer their level of agreement with ten items about the impact on patients of accessing their online records. Employing 4-level Likert items, we included the following response options: “disagree,” “somewhat disagree,” “somewhat agree,” “agree.” All closed ended questions included “don’t know” options.

Part Two of the survey requested participants to “please think about how your practice will be affected or already is affected if your patients have full online access to their GP health record.” Employing the same 4-level Likert scale, participants were requested to offer their opinions about nine survey items on the impact to GPs and their practices of patient access. This was followed by a question requesting participants to rate the legal risk of actions being taken against them as a result of online record access. Participants were offered three options: “decrease my risk of having legal action taken against me,” “increase my risk of having legal action taken against me,” or “neither decrease nor increase my risk.” All of the above closed ended questions also included “don’t know” options. A final question asked, “Are you aware that, since April 2019, the GP contract in England has required GP surgeries to offer patients full access to all prospective data on their GP health record?” Participants were requested to answer this question using only binary ‘yes’ or ‘no’ options.

Part One and Part Two of the questionnaire also included four open comment questions. Responses to these questions have now been analysed and published.[22].

The survey closed by requesting demographic information including participant gender, and age. Participants were also asked whether they were willing to be contacted to participate in a follow-up online survey in 2023.

### Data management and analysis

We used descriptive statistics to examine physicians’ characteristics and experiences with their opinions about ORA. In our analysis, responses were collapsed into positive (for “somewhat agree” or “agree” responses) versus negative (for “somewhat disagree” or “disagree”) opinions. The chi-square test of independence and Fisher’s exact test were performed to check the associations of physicians’ experiences and opinions about the impact of ORA with age, gender and working hours per week. The latter analysis was included as previous surveys in the US suggest clinicians fear ORA will exacerbate already heavy workloads, contributing to greater time in writing documentation [10,11,13]; we therefore surmised that GPs who worked longer hours would be more sceptical about the practice. We completed all analysis using SAS software version 9.4 (SAS Institute Inc., Cary, N.C.). Figures were created with Datawrapper.

## Results

### Respondent characteristics

Of the 720 who received email and homepage invitations, 601 opened the email invite, and 102 clicked on the survey link with 63 completing the survey (response rate: 63/720, 9%); the remainder (337) accessed and completed the survey via their homepage (337/2072, 16%). Of the 400 GPs who responded more were male (57%), and 85% were aged 40 or older. Respondents were from all regions of England. Most of our respondents worked between 21 and 40 hours per week (58%, 230/400). See Table 1.

**Table 1.** Characteristics of the respondents and their practices.

| Characteristic | Sample<br>(*n* = 400) | GMC Register<br>(*n* = 59,001) | Chi-square |
| --- | --- | --- | --- |
| **Gender\*, *n* (%)** | - |  | 26.39 |
| Female | 161 (40.25%) | 32,171 (54.53%) |  |
| Male | 227 (56.75%) | 26,830 (45.47%) |  |
| Prefer not to disclose | 12 (3%) | - |  |
| **Age\*, *n* (%)** |  |  | 70.91 |
| 25 – 29 | - | 77 (0.13%) |  |
| 30 – 39 | 61 (15.25%) | 14,558 (24.67%) |  |
| 40 – 49 | 196 (49%) | 18,518 (31.39%) |  |
| 50 – 59 | 99 (24.75%) | 13,816 (23.42%) |  |
| 60 + | 44 (11%) | 12,032 (20.39%) |  |
| **Hours worked per week, m (SD)** |  |  |  |
| 0 – 20, n (%) | 65 (16.25%) | - |  |
| 21 – 40, | 230 (57.5%) | - |  |
| 41 + | 105 (26.25%) | - |  |
| **Location of practice, n (%)** |  |  | **3.16** |
| London | 64 (16%) | 10,150 (17.2%) |  |
| South West | 45 (11.25%) | 8,091 (13.71%) |  |
| South East | 64 (16%) | 9,305 (15.77%) |  |
| East & West Midlands | 77 (19.25%) | 10,274 (17.41%) |  |
| East of England | 40 (10%) | 5,588 (9.47%) |  |
| North East & Yorkshire and Humber | 58 (14.5%) | 8,259 (14%) |  |
| North West | 52 (13%) | 7,335 (12.43%) |  |

\*Statistical significance found on age (df=2) and gender (df=1) by comparing the calculated Chi-square values with critical values with the significant level at 0.05

Our participants varied from those registered with the General Medical Council (GMC) in March 2022. There were more male GPs in our sample than those in the GMC registry (57% Vs 45%). Our respondents were also older than those in the registry; 85% Vs 75% aged 40 and above. Our sample was representative of the seven English regions. Since the GMC does not collect the number of hours worked per week it was not possible to compare participants on this metric.

### Experiences and Opinions about Impact on Patients

Around a quarter of surveyed GPs (28%, 111/400) estimated that 51 to 100% of their patients currently had access to their full online health records. A further 12% (50/400) reported “none” with 21% (83/400) reporting they “don’t know” what percentage were offered online access (Fig 1). Six in ten participants (60%, 240/400) believed that if patients were offered access to their free text entries only 50% or fewer would read them (see Fig 1).

**Figure 1.**
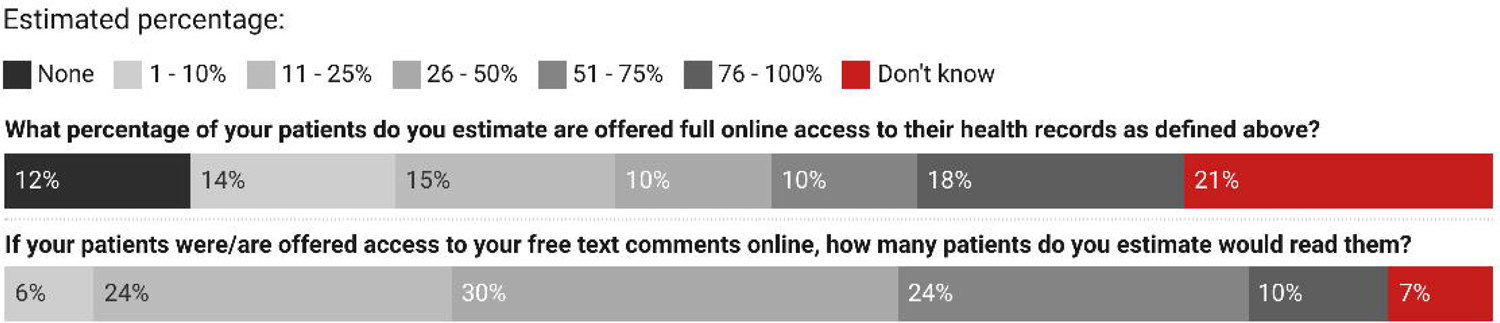
Estimated percentage of patients offered full access to health records and of patients reading them.

Approximately nine in ten participants (91%, 364/400) somewhat agreed or agreed that after obtaining full online access, a majority of patients would “worry more” with 85% (338/400) believing most patients would “find their GP health records more confusing than helpful” (see Table 2). Similarly, 95% (381/400) somewhat agreed or agreed that after full online access, a majority of patients would “contact me or my practice with questions about their health record.”

**Table 2.**
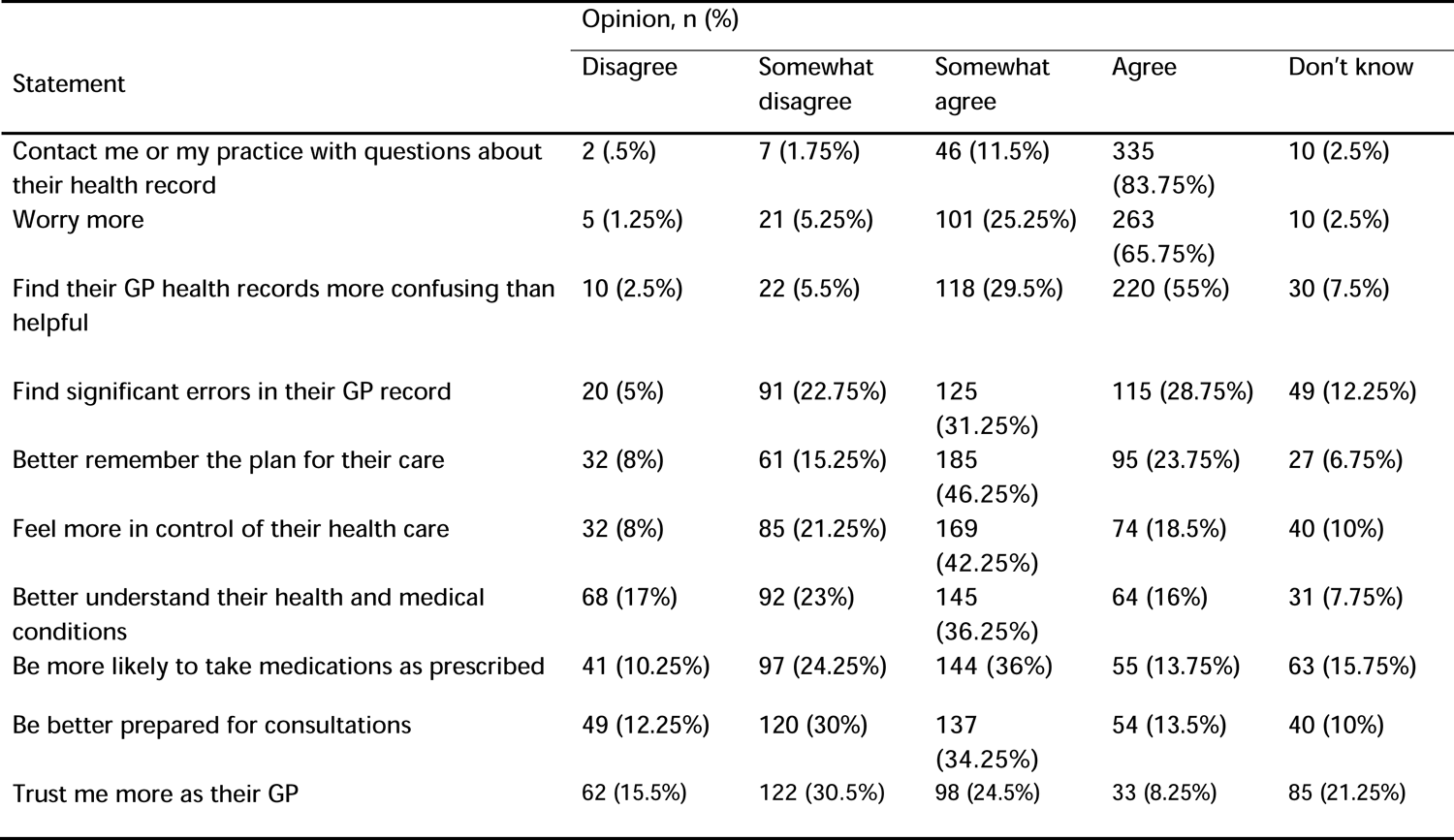
GPs’ Experiences and Opinions about the Impact of Online Record Access on Patients.

| Statement | Opinion, n (%) |  |  |  |  |
| --- | --- | --- | --- | --- | --- |
|  | Disagree | Somewhat disagree | Somewhat agree | Agree | Don't know |
| Contact me or my practice with questions about their health record | 2 (.5%) | 7 (1.75%) | 46 (11.5%) | 335 (83.75%) | 10 (2.5%) |
| Worry more | 5 (1.25%) | 21 (5.25%) | 101 (25.25%) | 263 (65.75%) | 10 (2.5%) |
| Find their GP health records more confusing than helpful | 10 (2.5%) | 22 (5.5%) | 118 (29.5%) | 220 (55%) | 30 (7.5%) |
| Find significant errors in their GP record | 20 (5%) | 91 (22.75%) | 125 (31.25%) | 115 (28.75%) | 49 (12.25%) |
| Better remember the plan for their care | 32 (8%) | 61 (15.25%) | 185 (46.25%) | 95 (23.75%) | 27 (6.75%) |
| Feel more in control of their health care | 32 (8%) | 85 (21.25%) | 169 (42.25%) | 74 (18.5%) | 40 (10%) |
| Better understand their health and medical conditions | 68 (17%) | 92 (23%) | 145 (36.25%) | 64 (16%) | 31 (7.75%) |
| Be more likely to take medications as prescribed | 41 (10.25%) | 97 (24.25%) | 144 (36%) | 55 (13.75%) | 63 (15.75%) |
| Be better prepared for consultations | 49 (12.25%) | 120 (30%) | 137 (34.25%) | 54 (13.5%) | 40 (10%) |
| Trust me more as their GP | 62 (15.5%) | 122 (30.5%) | 98 (24.5%) | 33 (8.25%) | 85 (21.25%) |

In contrast, 70% (280/400) somewhat agreed or agreed that a majority of patients would “better remember the plan for their care,” with 61% (243/400) believing patients would “Feel more in control of their health care.” Similarly, 60% (240/400) somewhat agreed or agreed that a majority of patients would “find significant errors in their GP record.” Around half of those surveyed (52%, 209/400) somewhat agreed or agreed that a majority of patients would “better understand their health and medical conditions” after accessing their online records, or “be more likely to take their medications as prescribed” (50%, 199/400). A similar proportion (48%, 191/400) somewhat agreed or agreed that after obtaining online access, a majority would “be better prepared for consultations.”

When asked about the influence of access on trust, more than four in ten respondents (46%, 184/400) believed ORA would have a negative impact.

### Experiences and Opinions about Impact on GPs

More than eight in ten participants (85%, 338/400) reported they were “aware that, since April 2019, the GP contract in England has required GP surgeries to offer patients full online access to all prospective data on their GP health record.” Almost nine in ten respondents (89%, 357/400) somewhat agreed or agreed that “I will/already spend significantly more time addressing patient questions outside of consultations” as a result of patient access to their online health records (see Table 3). Similarly, 81% (322/400) somewhat agreed or agreed that “my consultations will take/already take significantly longer.” In addition, 72% (289/400) somewhat agreed or agreed that “I will be/already am less candid in my documentation.” A minority of respondents (18%, 72/400) agreed or somewhat agreed that “medical care will be/is delivered more efficiently” after patient access to their GP health record. Around two in three respondents (64%, 255/400) disagreed or somewhat disagreed that “patient satisfaction will improve/has already improved.” Asked whether patient care will be/is safer as a result of patient online record access, 57% (227/400) disagreed or somewhat disagreed; 19% (74/400) responded “don’t know.” In addition, 58% (230/400) somewhat agreed or agreed that “patients who read their GP record will be/already have been offended.” Nearly two thirds of respondents (62% 246/400) believed patient online access would “increase my risk of having legal action taken against me.” See Fig 2.

**Figure 2.**
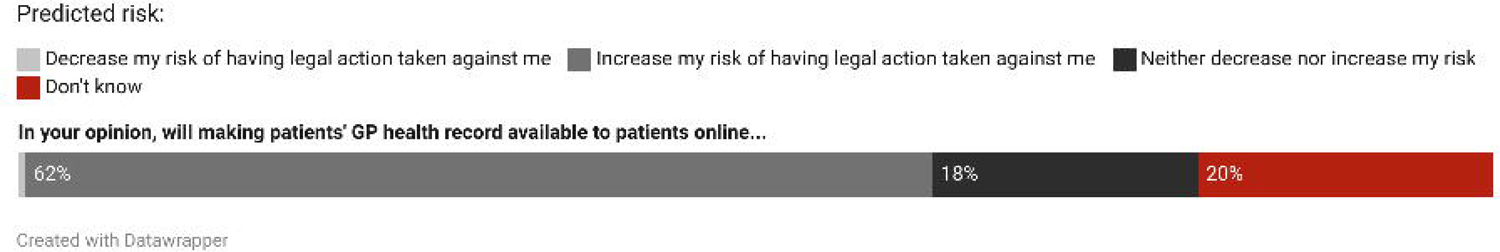
Predicted risk of legal action.

**Table 3.**
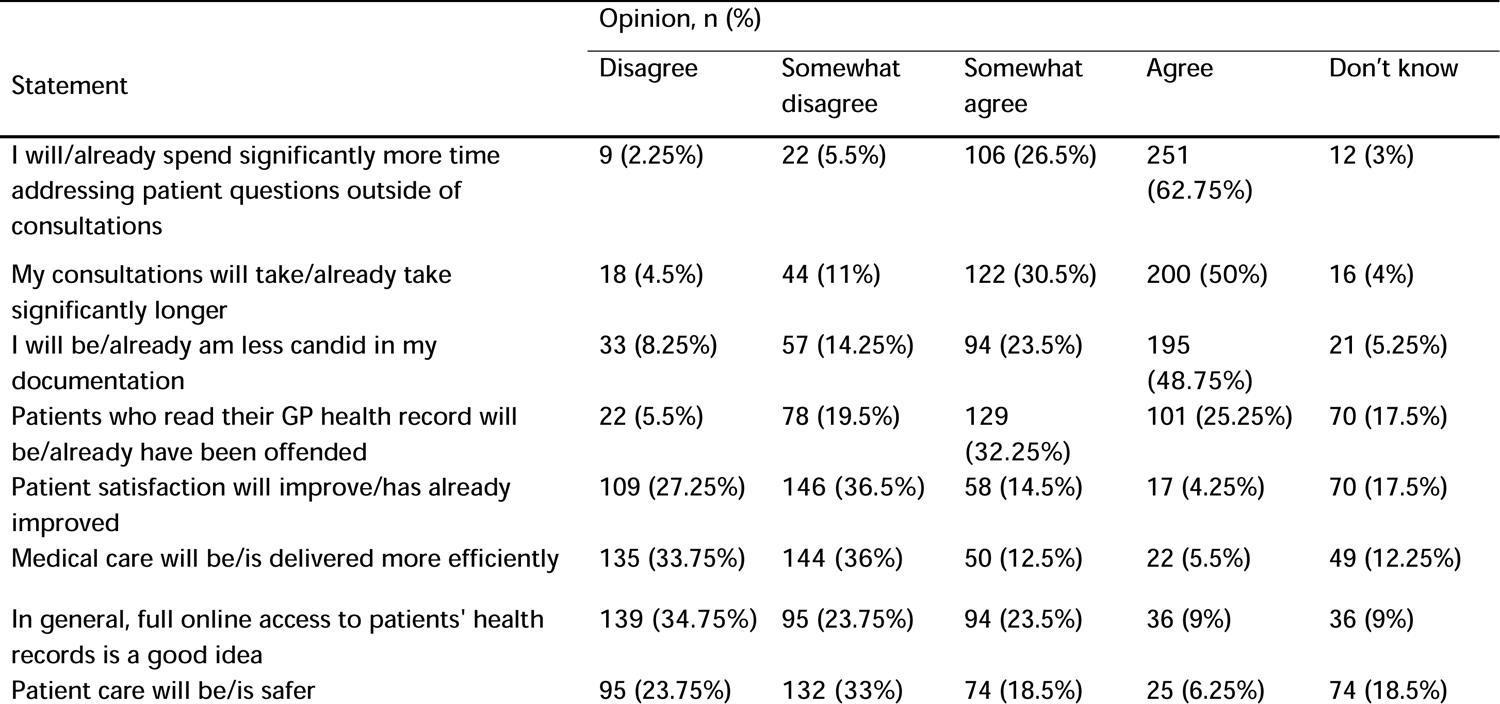
GPs’ Experiences and Opinions about the Impact of Patient Online Health Records Access on their Practice.

Finally, a third of respondents (33%, 130/400) somewhat agreed or agreed that patient access to their online records was a “good idea.”

## Discussion

### Summary of major findings

This is the largest survey conducted into the experiences and opinions of GPs in England about patient online record access. Most respondents were aware that the GP contract in England required surgeries to offer patients full online access on request. By March 2022, a quarter of GPs in our study reported enabling online records access for most of the majority of their patients. Only a third of surveyed GPs believed offering patients access to their online records was a good idea.

The overwhelming majority of surveyed GPs believed patients would worry more after accessing their records with a similar proportion (85%) believing most would find their records more confusing than helpful. However, our findings also revealed GPs believed there were benefits to patients from access. The majority of GPs believed access would improve patient recall about their care plan, enhance patients’ sense of control over their care, and help patients to identify significant errors in their record. In addition, approximately half of those surveyed believed access would help patients better understand their health and medical conditions, better adhere to their medications, and to be better prepared for consultations.

Despite experienced and anticipated benefits to patients, GPs worried about the work burdens of ORA on their practices. More than nine in ten surveyed GPs believed patient online access would lead to increased patient contact. Most GPs also believed consultations will/already take longer as a result, with more time spent answering questions outside of consultations. Perhaps as a result, seven in ten GPs did not agree access would increase care efficiency. The majority were also sceptical that patient satisfaction or safety would improve as a result of patient access. Finally, more than six in ten GPs believed the risk of patients taking legal action against them would increase after online record access.

### Comparison with other studies

Our survey supports recent qualitative research in England which shows clinicians express partial ambivalence, and scepticism about the impact of ORA on patients.[23,24] Moreover, strikingly similar views have also been reported cross-culturally in countries where access is now advanced. For example, multiple surveys in the U.S. and Sweden reveal a majority of clinicians, especially those with limited experience of ORA, expressed hesitancy or resistance to the practice.[12,25–27] Relatedly, most respondents reported awareness that the GP contract committed practices to offer patients full online access by request, yet only a minority reported offering most patients in their practice online access to their records.

Professional reluctance to raise awareness about access is also found in other countries. For example, in a survey of doctors in the US, even after opening notes to patients, 78% (n=620) admitted that they did not encourage patients to read their documentation.[28] Indeed, in a recent qualitative study among patients in England, participants reported online access should be better promoted.[18]

Our results also echo studies in other countries which show doctors anticipate patients will feel more in control of their care, and better remember their care plan after accessing their records.[10,11] Equally, our findings also resonate with research conducted in Sweden and the U.S. demonstrating most clinicians doubt patients’ ability to handle what they read, anticipating patients will worry more, and find their records more confusing than helpful.[10–12] Clinicians’ doubts tend to dissipate over time and with experience of ORA.[29] Notably, to date, studies consistently show the majority of patients who access their records – including those with chronic illnesses – describe multiple benefits with few reporting feeling ‘very confused’ or ‘more worried’ by what they read.[19,20,30–34]

Again, like previously published surveys in other countries, most of the English GPs we surveyed worried about access encroaching on their workload.[23,24] This theme is also predominant in survey findings in other countries. For example, in a recent U.S. study of 116 primary care physicians, 69% anticipated spending significantly more time addressing patients’ questions outside of consultations prior to patient access to open notes (that is, free text entries about patient visits); after implementation, only 8% reported having to do so.[29] Using objective measures of messaging—such as email volume—in 2012 a US survey by Delbanco *et al* found no significant changes in the 12 months before compared with the 12 months after open notes were implemented.[10] A more recent US survey led by DesRoches, among clinician respondents who had offered online patient access to open notes for at least 1 year, 86% (n=1112) reported that in the previous 12 months, patients contacted them less than monthly or never with questions related to their documentation.[28] However, other studies show that when it comes to online, or access to the full electronic health record there is potential for increased patient contact. In a systematic review in primary care settings, Mold et al found provision of online record access resulted in a moderate increase in e-mail traffic, but no change in telephone contact, with variable changes to face-to-face contact.[35] Another recent study in the US found that, after implementation of ORA, the number of messages sent by patients within the 6 hours after patients reviewed result doubled.[36]

More than half of surveyed GPs in our study believed patient access would negatively affect patient safety, a finding that was particularly noteworthy considering 60% (240/400) of GPs agreed that patients would find significant errors in their notes. Our respondents’ views contrast with multiple studies which suggest open notes might function as a safety mechanism,[37–39] a conclusion that is supported by systematic reviews and meta-analyses.[40,41] Studies show, with more eyes on the record, access may help patients and their families avoid delays and missed diagnoses by encouraging prompt follow up of tests, results, and referrals.[39,42]

GPs’ safety concerns may also have been driven by potential changes to documentation. Akin to recent qualitative studies among primary care staff in England,(21) and survey findings from other countries,(11,13,26) most GPs (72%, 279/400) in our survey reported they will be/already are less candid in their documentation as a result of patient online access. Such changes may be aimed at preventing patient anxiety or reducing anticipated patient contact, reducing litigation, or unintended offence. Nonetheless, after implementation of ORA, whether changes to documentation do, in fact, diminish the clinical value of documentation is unclear.[43] In the survey by DesRoches, 77% (n=188) of primary care physicians perceived no change in the value of their notes for other clinicians, however 26% (n=63) reported changing how they wrote differential diagnoses.[28] In addition, and although not determined in this survey, third party access or how to protect vulnerable patients, might also have been a safety concern for our respondents. Recent qualitative studies in England reported that primary care staff identified patient safeguarding including for at risk adults, such as those in coercive relationships, or among vulnerable young adults, as a leading concern.[23,24]

Finally, supporting recent qualitative research in England, a majority of GPs worried about elevated risks of litigation following ORA.[23] In the U.S. to date, we are aware of no medical malpractice cases arising as a result of patient access to their online records. If clinicians make changes that reduce the quality of documentation and this later leads to error, risks of litigation might increase. However, if patient access helps increase patient safety by reducing diagnostic delays or medical errors, this could reduce the risk of malpractice since these are the leading causes of claims.[44–47]

In summary: our survey was in line with recurrent themes in the growing body of international research into clinicians’ views about online record access. However, important contextual, including country-specific factors might have influenced our results. Compared with previously published clinician surveys, the present survey was administered in March 2022, during the COVID-19 pandemic and GP burnout may have contributed to increased cynicism. In the UK, successive governments have advocated a “digital first” model of primary care with ambitious short-term goals for transforming access to health advice, support, and treatment using digital online tools.[48,49] These policies, accelerated by the pandemic, may have exacerbated GPs’ concerns about work burdens, the rapid adoption of digital tools without adequate training, resources for implementation, or consideration to the possible negative consequences of these policies.[23]

### Implications of the findings

Our findings suggest GPs in England share many similar concerns with their counterparts in other countries where online access to records is now well established. Although few studies have explored patients’ experiences with online access in England,[14] we cannot help but observe a trend toward contrastive views between clinicians and patients. Combined, these findings suggest patients in England may be vulnerable to negative stereotyping with regard to their capacity to understand, and emotionally cope with, reading their own health information.[27] Medical ethicists have argued that unfair stereotyping may be used to justify exclusions which further impede patients’ ability to engage in their own care, forfeiting important opportunities to benefit from accessing their documentation.[27,50–52] The current study underscores the importance of exploring patients’ experiences in England with online record access.

Notwithstanding, GPs in our survey did perceive many benefits to patients. This is an important finding, given a growing body of research in other countries that patients feel more engaged, more in control of their care, and better understanding their care plan as a result of access to their clinical documentation.[19,20,30,33,34] Moreover, in these surveys, as a result of ORA, many patients report greater trust in their clinicians,[42,53] a greater sense of teamwork,[42] and doing a better job taking their medications.[33]

However, as with surveys in other countries, many GPs believed access would increase work burdens, and contact from patients. Again, studies in other countries suggest that with practice these fears may not materialise. Our study highlights the importance of supporting GPs and their staff to become better prepared for talking about and writing documentation that patients will now read.[54] Equally, patients will require guidance to optimise the benefits, and minimise risks. Guidance materials should be aimed at supporting GPs and patients to better partner with each other and to promote engagement with care plans while raising awareness about GPs’ work burdens.

### Strengths and limitations

This is the largest survey conducted in the UK on doctors’ views about patient access to their online health records. Given the ongoing changes to default online patient record access, the survey is timely. However, the study has several limitations relating to the use of a non-probability sample. Although we strove to stratify the sample, as far as possible according to geographical location, gender, and age, our respondents were restricted to those GPs who use Doctors.net.uk, and who used the service during the administration of the survey. It is not possible to infer that our sample was representative of the opinions of GPs in England. Furthermore, the decision to complete the survey may have been influenced by responder biases such as acquiescence biases or prior enthusiasm or scepticism about the topic which might have affected findings.

We recommend future studies conduct more in-depth analyses of GPs’ ongoing experiences, and opinions about ORA. To that end, a follow-up panel survey among participants who agreed to be contacted is planned. Few studies in England have explored patients’ experiences with accessing their online health records. Whether patients in England also accrue the same benefits as patients in other countries remains to be seen. Conceivably, there may be differences in documentation practices or in health literacy between countries, and we strongly recommend survey research in England explore the views of patients and their families with this practice innovation. As with other countries, further studies are also needed to explore objective changes to documentation as a result of ORA,[43,55] and to investigate the potential impact to workflow among clinicians following patient access.[36]

## Conclusions

Most GPs in this England-wide survey agreed there were multiple benefits to patients of accessing their online health records. Nonetheless, like clinicians in other countries, a majority of surveyed GPs believed patients would worry more and find their records more confusing than helpful, with increased contact with patients, and added work burdens. We emphasise that studies of patients’ experiences in diverse countries question the robustness of this perspective; however, it will be important for ongoing studies in the UK to evaluate and continue to assess both GPs’ and patients’ experiences with access. Notwithstanding, in England, patients’ online access to their GPs’ records is here to stay. In the coming months, it will be crucial for GPs, primary care staff, and patients, to adapt to this radical change in practice [54]. (See Box 1).

### Box 1.

#### Key questions and findings

##### What is already known about this topic

Ø In a growing number of countries worldwide patients are offered online access to their full health record.
Ø Since November 2022, NHS England has committed practices to offer new patients full online access to records.
Ø A growing body of research from countries (where patient access is advanced) shows patients derive multiple benefits from accessing their online records including greater engagement in their care, better understanding of, and strengthened recall about their treatment plan.

##### What this study adds

Ø Only a third of surveyed GPs believed patient access to their online records was a good idea. GPs believed access would incur both negative and positive effects on patient care.
Ø Around nine in ten believed a majority of patients would worry more, with eight in ten believing a majority of patients would find their GP records more confusing than helpful.
Ø In contrast, most GPs believed a majority of patients would find significant errors in their records, and that patients would better remember their care plan and feel more in control of their care as a result of online record access.
Ø Most GPs believed access would exacerbate work burdens and inefficiencies. More than eight in ten GPs believed access would lead to more time addressing patients’ questions outside of consultations and that consultations will take significantly longer. Nearly two thirds of GPs believed access would increase their risk of legal action against them.

## Supporting Information

## Data Availability

Data provided in Supplement 2.

## Acknowledgments

The authors acknowledge Jan Walker for permission to adapt survey items from an earlier study of open notes in the U.S.

