## Supplement 1 for "Patients Accessing their Online Records in England: A Survey of General Practitioners’ Experiences and Opinions"

**Medical Records Access Survey for GPs**

L1

Researcher names: Dr Catherine DesRoches and Dr Charlotte Blease
Contact: Beth Israel Deaconess Medical Center
Title of Research Project: A survey of UK GPs' Experiences and Opinions about Patients' Online Access to their Full Health Records

You are invited to participate in a research study conducted by Beth Israel Deaconess Medical Center, Harvard Medical School on patients' access to their online health records. If you decide to take part in this study you will be asked to answer questions on your experience, and opinions about offering patients' full online access to their health records. You are invited to participate because we are interested in your insights as a GP on offering patients' access to their online health records and the impact/potential impact you believe this might have on your practice. Participants who respond will also be invited to offer their opinions about patients' access to their online records, 12 months later. Therefore, if you decide to participate, we request that you be willing to respond to a second 5 minute survey at a later date.

This study has received ethical approval from Beth Israel Deaconess Medical Center Protocol #: 2021P000626. Taking part is entirely voluntary and before taking part you will be asked to complete a consent form. All of your answers will be completely anonymous. In the survey we will not ask for your name or any personal details about you such as contact details. The information you provide will be identified by a numerical code. The survey will be managed by Doctors.net.uk This survey is funded by FORTE, the Swedish Research Council for Health, Working Life and Welfare.

The study will begin with some questions asking for your awareness and opinions about patient access to their online health records in primary care. This will be followed by questions asking for your opinions/experiences about how online access might affect patient care, and your practice. The survey will close with some brief questions about your current employment and demographic questions (your age, gender, year when you started practicing medicine ). Most questions will ask you to answer by choosing from options, some questions will ask you to insert a number (e.g. year when you started practicing as a GP, number of hours worked per week). The survey also includes some optional open comment boxes.

The questionnaire will take no longer than 5 minutes to complete and all participants completing the survey in full will receive 1,000 eSR points

If at any time you change your mind and decide to abort the study you can do so. Both partial and completed responses will be saved; but partial responses will later be deleted for the purposes of data analysis.

If you decide to take part in the study, you will help improve understanding of the medical community's views about sharing online access to patients' health records. At the end of the study all of the completed surveys received will be used in the publication of a paper in a scientific journal. Details published will be anonymous

You can contact the Principal Investigator is Dr Catherine DesRoches and the Co-Investigator is Dr Charlotte Blease at any time if you have any concerns or questions about the study Contact details are:. If you change your mind and would like to withdraw . Contact details are: If you wish to be kept informed about the results of this project you can contact Dr. DesRoches or Dr. Blease directly.

Contact details will be provided again at the end of the questionnaire.

If you decide to take part in this study, we appreciate your time and thank you.

Consent : To take part in the survey, please confirm you agree with all the following statements.

- I confirm that I have read and understand the information supplied above for the study (1)
- I understand that my participation is voluntary and that I am free to withdraw from the study at any time without giving reason (2)
- I agree to the use of anonymised quotes in the reporting of the findings (3)
- I understand that all the study data and documentation whether hardcopies or electronic will be kept for up to 10 years and will be disposed of securely if it is confirmed that they are no longer required (4)
- Anonymised data may be accessed by staff at Beth Israel Deaconess Medical Center and regulatory bodies for audit and monitoring purposes (5)
- I agree to take part in the above survey (6)
- No (7)

info

The first section of the survey is a set of screening questions to determine if the survey will be relevant to you. There are a maximum of 3 screening questions.

S1 - Role

Please select your role from the list below:

- GP (1)
- Hospital specialist (2)
- Other (3)

S2 - Country

Whereabouts are you currently practising?

- England (1)
- Wales (2)
- Scotland (3)
- Northern Ireland (4)
- Republic of Ireland (5)
- Practicing elsewhere (6)
- Retired (7)

S3 – Region

Where are you currently practicing?

Please note that for England, we are referring to the standard regions rather than the NHS regions: https://en.wikipedia.org/wiki/Regions_of_England

- London (1)
- South West (2)
- South East (3)
- West Midlands (4)
- East Midlands (5)
- East of England (6)
- Yorkshire and Humber (7)
- North East (8)
- North West (9)

Part 1: Impact to Patients of online health records access

The following questions ask for your understanding, experiences, and opinions about offering patients full online access to their GP health records including the potential impact on patients' care.

By "full online access" we mean all information on the electronic record from the date the patient requested access that is visible to GPs including the patient's allergies, immunizations, letters, medications lists, test/lab results, problem lists, and the free text comments written by clinicians.

Q1

What percentage of your patients do you estimate are offered full online access to their health records as defined above?

- None (1)
- 1-10% (2)
- 11-25% (3)
- 26-50% (4)
- 51-75% (5)
- 76-100% (6)
- Don't know (7)

Q1a

Do you offer access to any of these?

Please select all options that apply.

Transactional functions

- Appointment booking/cancelling (1)
- Medication requests (2)
- Referral information (3)
- Online messaging (patient initiated) (4)
- Questionnaire completion (5)
- None of these (6)

Q1ai

Access to information in the record

- Allergies (7)
- Immunisations (8)
- Letters (e.g. hospital letters) (9)
- Medications list (10)
- Test/lab results (11)
- Problems list (12)
- Free text comments written by clinicians (13)
- None of these (14)

Q1c

Please add any additional comments you might have about the information your patients can access.

Optional

info

For the following questions please assume that all your patients have been granted full online access to their GP Health record. By "full online access" we mean all information on the electronic record from the date the patient requested access that is visible to GPs including the patient's allergies, immunizations, letters, medications lists, test/lab results, problem lists, and the free text comments written by clinicians.

Q2

If your patients were/are offered access to your free text comments online, how many patients do you estimate would read them?

- 0-10% (1)
- 11-25% (2)
- 26-50% (3)
- 51-75% (4)
- 76-100% (5)
- Don't know (6)

Q3

We are interested in your opinions about the effect on patients of reading GP health records online, even if none of your patients have requested access. Please indicate how strongly you agree or disagree with the statements below.

 Among my patients who read their full GP health record online a majority will:

|  | Disagree (1) | Somewhat disagree (2) | Somewhat agree (3) | Agree (4) | Don't know (5) |
| --- | --- | --- | --- | --- | --- |
| Better understand their health and medical conditions (1) |  |  |  |  |  |
| Worry more (2) |  |  |  |  |  |
| Better remember the plan for their care (3) |  |  |  |  |  |
| Be more likely to take medications as prescribed (4) |  |  |  |  |  |
| Find significant errors in their GP record (5) |  |  |  |  |  |
| Feel more in control of their health care (6) |  |  |  |  |  |
| Be better prepared for consultations (7) |  |  |  |  |  |
| Trust me more as their GP (8) |  |  |  |  |  |
| Contact me or my practice with questions about their health record (9) |  |  |  |  |  |
| Find their GP health records more confusing than helpful (10) |  |  |  |  |  |

Q4

Do you have any additional comments about the impact to patients accessing their full GP health record online?

Optional

info

Part 2: Impact of Patient online health records access on my practice

In this section, please think about how your practice will be affected or already is affected if your patients have full online access to their GP health record.

Q5

|  | Disagree (1) | Somewhat disagree (2) |  | Somewhat agree (3) | Agree (4) | Don't know (5) |
| --- | --- | --- | --- | --- | --- | --- |
| My consultations will take/ already take significantly longer (1) |  |  |  |  |  |  |
| I will/already spend significantly more time addressing patient questions outside of consultations (2) |  |  |  |  |  |  |
| Patients who read their GP health record will be/already have been offended (3) |  |  |  |  |  |  |
| I will be/already am less candid in my documentation (4) |  |  |  |  |  |  |
| Medical care will be/is delivered more efficiently (5) |  |  |  |  |  |  |
| Patient satisfaction will improve/has already improved (6) |  |  |  |  |  |  |
| Patient care will be/is safer (7) |  |  |  |  |  |  |
| In general, full online access to patients' health records is a good idea (8) |  |  |  |  |  |  |

Q5a

|  | Decrease my risk of having legal action taken against me (1) | Increase my risk of having legal action taken against me (2) | Neither decrease nor increase my risk (3) | Don't know (4) |
| --- | --- | --- | --- | --- |
| In your opinion, will making patients' GP health record available to patients online: |  |  |  |  |

Q6

Do you have any additional comments about the impact of patient access to their full online health records on your practice?

Optional

Q7

Are you aware that, since April 2019, the GP contract in England has required GP surgeries to offer patients full access to all prospective data on their GP health record?

- Yes (1)
- No (2)

D1 - Hours worked

On average, how many hours per week do you work?

Please do not include time spent supervising trainees.

| (1) | ______________________________ | hours/week |
| --- | --- | --- |

D2 - Gender

Your gender

- Male (1)
- Female (2)
- Not listed above: Please specify: (3)____________
- Prefer not to say (4)

D3 - Age

Your age

- 29 years or younger (1)
- 30-39 (2)
- 40-49 (3)
- 50-59 (4)
- 60 or older (5)

F1 - Feedback question

Do you have any additional comments, anecdotes, and/or expectations to share about patients' online access to their full online health record including clinicians' free text entries?

Optional

F2

Would you be willing to be contacted to take part in a follow up online survey in 2023 on the impact of patient access to their medical records?

- Yes (1)
- No (2)
